# Safety and Efficacy of Pemivibart, a Long-Acting Monoclonal Antibody, for Prevention of Symptomatic COVID-19: Interim Results From the CANOPY Clinical Trial

**DOI:** 10.1101/2024.11.11.24317127

**Authors:** Cameron R. Wolfe, Jonathan Cohen, Kathryn Mahoney, Anna Holmes, Natalia Betancourt, Deepali Gupta, Kazima Tosh, Kristin Narayan, Ed Campanaro, Chloe Katz, Anne-Marie Phelan, Ilker Yalcin, Mark Wingertzahn, Pamela Hawn, Pete Schmidt, Yong Li, Myra Popejoy, the CANOPY Study Group

## Abstract

**Background:** Pemivibart received emergency-use authorization for prevention of symptomatic COVID-19 in moderate-to-severe immunocompromised individuals based on immunobridging analysis in the phase 3 CANOPY trial. We report an interim analysis of safety and efficacy of pemivibart in individuals with (cohort A) or without (cohort B) significant immunocompromise over a contemporary variant landscape.

**Methods:** Eligible participants (aged ≥18 years; SARS-CoV-2-negative) received 2 intravenous 4500-mg pemivibart infusions (cohort A) or received blinded pemivibart or placebo (2:1, cohort B) 90 days apart. Safety was a primary endpoint. Composite incidence of reverse transcription-polymerase chain reaction (RT-PCR)-confirmed symptomatic COVID-19, COVID-19 hospitalization, or all-cause mortality was evaluated through month 6 (cohort A) and month 12 (cohort B).

**Results:** In September-November 2023, 306 participants with immunocompromise received pemivibart in cohort A; 317 received pemivibart and 162 received placebo in cohort B. The most common study drug-related adverse event was infusion-related reactions (cohort A: 11/306 [3.6%]; cohort B: 7/317 [2.2%, pemivibart] and 0/162 [placebo]). Four of 623 (0.6%) participants who received pemivibart experienced anaphylactic reactions (2 non-serious; 2 serious) within 24 hours of dosing. In cohort A, the composite COVID-19 endpoint incidence through month 6 (day 180) was 11/298 (3.7%; 2 deaths [suicide and unknown cause]) in participants who received a first full dose of pemivibart. In cohort B, the composite COVID-19 endpoint incidence through month 6 was 6/317 (1.9%) in participants in the pemivibart group and 19/160 (11.9%) in the placebo group, representing an 84.1% standardized relative risk reduction (RRR) (95% CI, 60.9-93.5; nominal *P*<.0001) for pemivibart. Through month 12, 15/317 (4.7%; 1 death [cardiac failure]) and 29/160 (18.1%) pemivibart and placebo participants met the composite clinical endpoint, respectively demonstrating a 73.9% standardized RRR (95% CI, 52.8-85.6; nominal *P*<.0001).

**Conclusions:** Pemivibart provided pre-exposure prophylactic efficacy against COVID-19 and was well-tolerated by most participants with or without significant immunocompromise.

Anaphylaxis was an important safety risk.

**Clinical Trials Registration:** NCT06039449

**Key points:** Pre-exposure prophylactic administration of 2 doses of pemivibart approximately 90 days apart was generally well-tolerated and provided protection against symptomatic COVID-19 through 6 months in individuals with immunocompromise and 12 months in individuals without immunocompromise respectively

## INTRODUCTION

Despite vaccine availability, SARS-CoV-2 continues to cause substantial morbidity from COVID-19 in the US and worldwide[1,2]. Immunocompromised individuals remain at high risk for severe disease, hospitalization, and death[3–6], with suboptimal vaccination response, waning vaccine effectiveness, and decreased vaccine uptake contributing to less protection[7–9].

Monoclonal antibodies (mAbs) may provide immediate, durable protection against symptomatic COVID-19 with reliable serum virus neutralizing antibody (sVNA) titers that do not depend on an intact immune system[10,11]. Several mAbs received emergency-use authorization (EUA) for COVID-19 prevention during the 2021-2022 pandemic[12–14]; however, all showed reduced activity to predominant circulating Omicron sublineages by late 2022 and were deauthorized[15], resulting in a substantial gap in preventive care for immunocompromised people[11].

Pemivibart is a half-life-extended human immunoglobulin G1 mAb engineered from adintrevimab, a mAb that demonstrated efficacy against symptomatic COVID-19 caused primarily by the Delta variant but subsequently lacked activity against Omicron[16,17]. Pemivibart functions as a human angiotensin-converting enzyme 2 competitor targeting an epitope on the spike glycoprotein receptor-binding domain of SARS-CoV-2[16–19]. In vitro, pemivibart has demonstrated neutralizing activity against multiple variants of SARS-CoV-2, including ancestral pre-Omicron sublineages (eg, D614G, Delta [B.1.617.2]) and Omicron sublineages (eg, JN.1, KP.3, KP.3.1.1, LB.1)[20]. In a phase 1 study, single intravenous (IV) doses of pemivibart ≤4500 mg were well-tolerated by healthy adults, with no drug-related serious adverse events (AEs)[21].

Pemivibart was granted EUA in the US in March 2024 for prevention of symptomatic COVID-19 in certain adults and adolescents (aged ≥12 years; weighing >40 kg) with moderate-to-severe immunocompromise who are unlikely to mount adequate immune response to COVID-19 vaccination[20,22]. The EUA was based on safety and immunobridging data using calculated sVNA titers of pemivibart against relevant SARS-CoV-2 variants from participants in CANOPY (NCT06039449[23]), a phase 3 clinical trial that evaluated pemivibart as pre-exposure prophylaxis against COVID-19 in 2 cohorts, individuals with or without significant immunocompromise.

Here we report data from analysis of safety, tolerability, clinical efficacy, pharmacokinetics (PK), and immunogenicity of pemivibart in CANOPY spanning a contemporary variant landscape.

## METHODS

### Study Design

CANOPY is a phase 3 study with 2 cohorts being conducted at 18 investigative sites in the US. Cohort A is an open-label, single-arm study that enrolled adults (aged ≥18 years) with significant immunocompromise (Table 1) to receive 2 single IV infusions of 4500-mg pemivibart 90 days apart. Cohort B is a randomized, triple-blind, placebo-controlled study that enrolled adults at risk of acquiring SARS-CoV-2 infection to receive either pemivibart or placebo (2:1, 90 days apart). The pemivibart dose level was fixed, with no adjustment required for body weight or renal or hepatic impairment. The trial included an up-to-14-day screening period and an efficacy and safety assessment period from time of initial dosing through month 12. The first participants in cohorts A and B were dosed on 14 and 8 September 2023, respectively. Safety outcomes are reported through the data cutoff (21 May 2024) when the last participant had completed ≥6 months of safety follow-up. Analysis of clinical efficacy of Cohort A is shown through 6 months and Cohort B through 12 months (data cutoff 07 Oct 2024), as these data were available at the time of this publication.

**Table 1.**
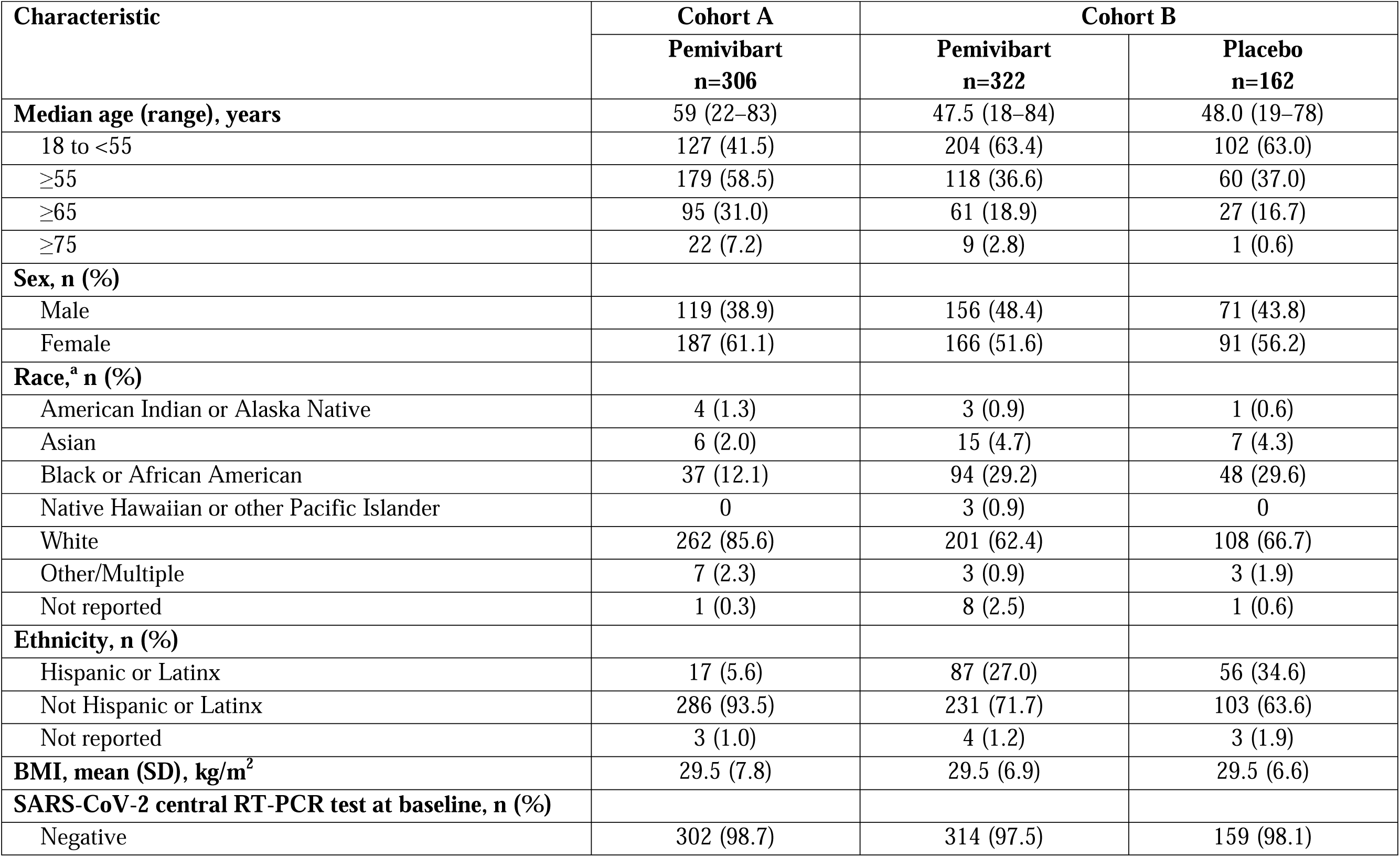
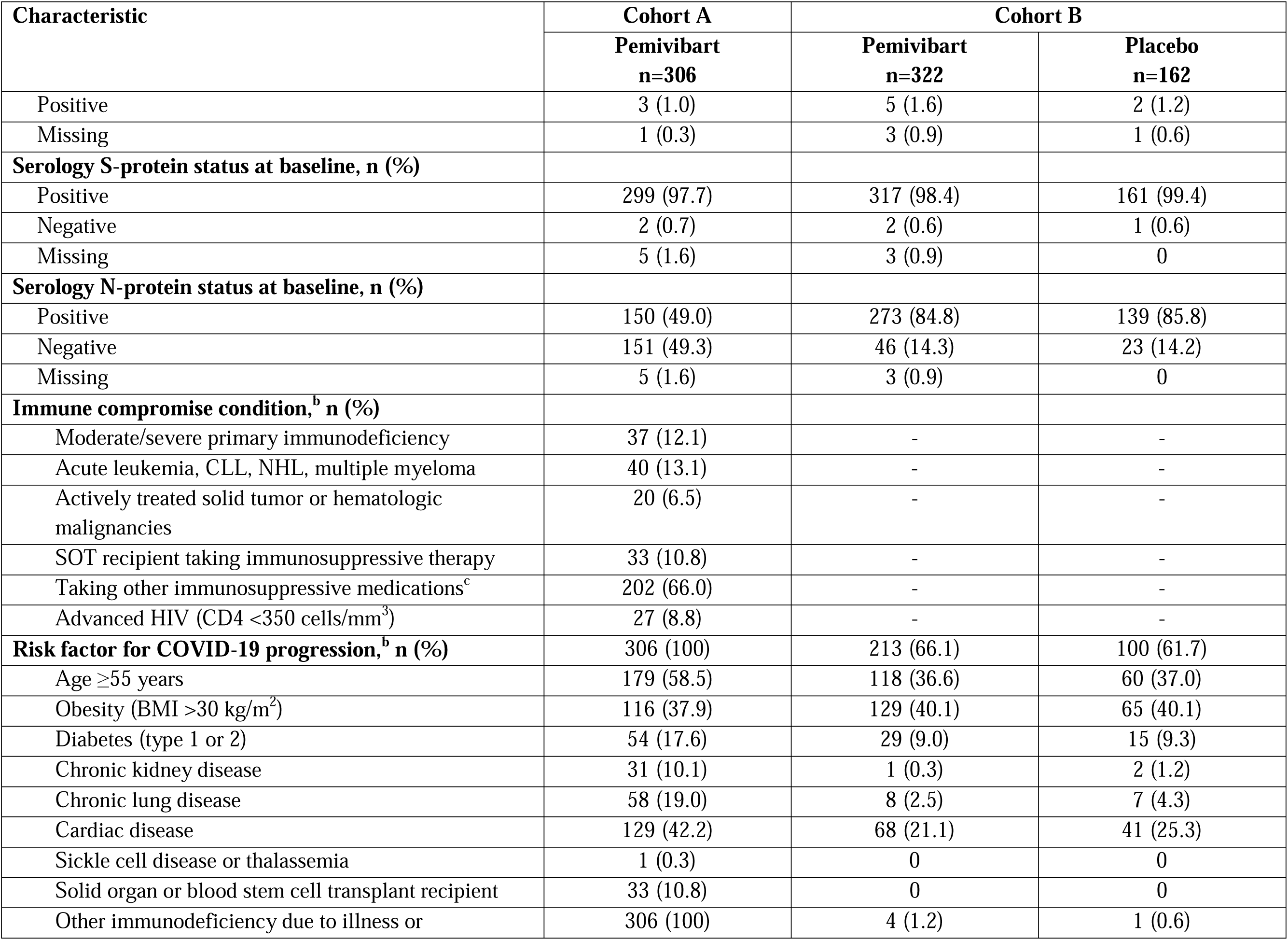
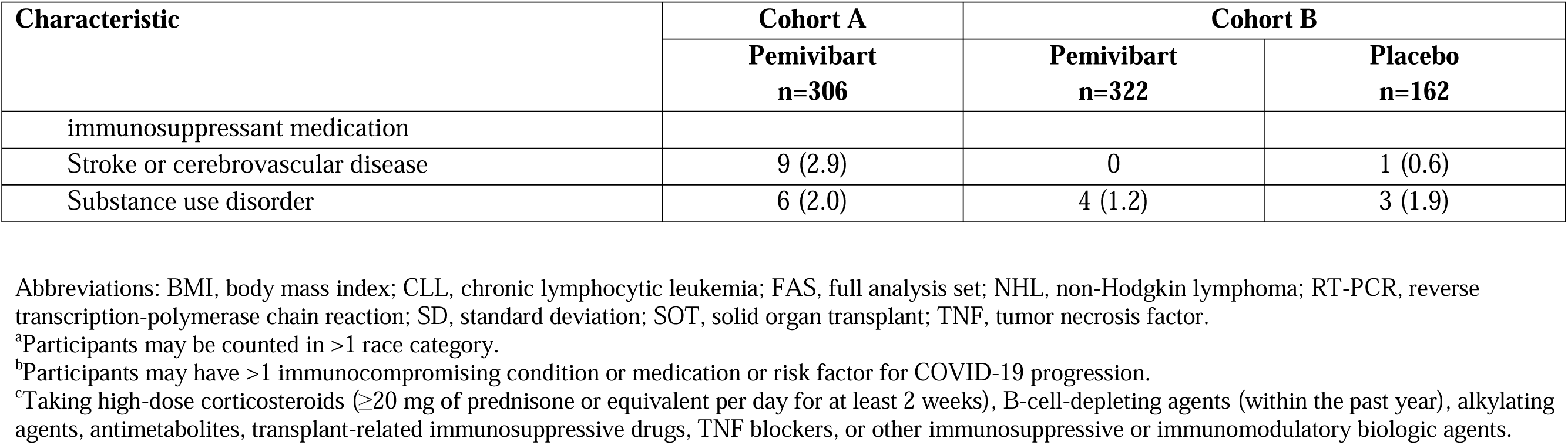
Demographic and Baseline Characteristics (Cohort A, Safety Analysis Set; Cohort B, FAS)

The trial is being conducted in accordance with the International Conference on Harmonization guideline on Good Clinical Practice, the principles of the Declaration of Helsinki, and all applicable regulations. Written informed consent was obtained from all participants. The protocol (including amendments) was approved by an institutional review board.

### Participants

Adults (≥18 years) who tested negative for SARS-CoV-2 infection by local antigen test or reverse transcription-polymerase chain reaction (RT-PCR) at screening and agreed to defer receipt of COVID-19 vaccinations or boosters for at least 28 days after the initial dose of study drug were eligible for enrollment. Participants eligible for cohort A had significant immune compromise (including active treatment for solid tumor or hematologic malignancies; acute leukemia, chronic lymphocytic leukemia, non-Hodgkin’s lymphoma, or multiple myeloma; solid-organ transplant (SOT) and taking immunosuppressive therapy; moderate or severe primary immunodeficiency; advanced or untreated HIV infection; active treatment with immunosuppressive drugs); those in cohort B were at risk of exposure to SARS-CoV-2 because of regular unmasked face-to-face interactions in indoor settings (eg, workplace, public transportation). Participants were excluded if they had received prior convalescent plasma or SARS-CoV-2 mAbs or had a SARS-CoV-2 infection ≤120 days before enrollment. COVID-19 vaccination was permitted ≥14 days before enrollment for cohort A and ≥120 days before randomization in cohort B. Full inclusion/exclusion criteria are provided in the Supplementary Materials.

### Cohort B Randomization and Blinding

For cohort B, randomization through an interactive response technology system was triple blinded for participants, investigators, and sponsor study team, with study drug assignments available to limited sponsor and site personnel for study drug preparation and supply activities. Randomization was stratified by age (12 to <55 years vs ≥55 years) and time since most recent SARS-CoV-2 infection or vaccination (120 days to ≤240 days vs >240 days or no prior infections or vaccinations). Between November-December 2023, an unblinded analysis was conducted for the EUA application by a select sponsor team. All sponsor personnel and designees working directly with the study sites remained blinded to individual treatment assignments per protocol.

### Assessments

Evaluation of the in vivo neutralizing activity of pemivibart against relevant SARS-CoV-2 variants[20] will be described in a separate publication. Nasopharyngeal swab and saliva samples collected from participants with self-reported symptoms of COVID-19-like illness were submitted for confirmatory testing at a central laboratory (PPD Global Central Lab, Highland Heights, KY). Blood samples for PK analysis and detection of antidrug antibodies (ADAs) were collected at specific timepoints through month 12 (see Supplementary Materials).

### Endpoints and Analysis Populations

Safety and tolerability of pemivibart was a primary objective for both cohorts. All participants were monitored for infusion-related or hypersensitivity reactions (IRRs/HSRs) for ≥1 hour postdose; treatment-emergent adverse events (TEAEs) were collected through the last follow-up visit before data cutoff. The safety analysis set included all participants who received any amount of study drug.

For cohort A, protection against symptomatic COVID-19 based on an immunobridging endpoint was a primary objective and was assessed in all participants who received a full dose of pemivibart and had at least 1 quantifiable serum concentration postdose (PK full analysis set [FAS]). For both cohorts, the clinical efficacy of pemivibart was assessed based on a composite endpoint of RT-PCR-confirmed symptomatic COVID-19 with onset of symptoms ≤14 days from positive sample collection or a COVID-19-related hospitalization or all-cause mortality through month 3 (day 90), following a single dose of pemivibart, and month 6 (day 180) and month 12 (day 365; cohort B only), following 2 doses of pemivibart administered 90 days apart. Clinical efficacy was evaluated in all participants in cohort A who received a full dose of pemivibart at initial dosing (FAS), and in all randomized participants in cohort B without current SARS-CoV-2 infection at baseline (confirmed by central laboratory RT-PCR) who received any amount of pemivibart or placebo (modified FAS; mFAS). Time to first event of the composite RT-PCR-confirmed symptomatic COVID-19 endpoint through month 6 (cohort A) and month 12 (cohort B) was summarized using the Kaplan-Meier method.

### Statistical Analysis

The statistical analysis plan for cohort A will be published in a separate manuscript. For cohort B there was no prespecified hypothesis testing. Because of unblinding that occurred for the EUA application, the statistical analysis plan was amended, and all cohort B efficacy endpoints were categorized as exploratory. Safety and exploratory endpoints were analyzed descriptively, and nominal *P*-values and 95% CIs were computed as applicable. Analysis of RT-PCR-confirmed symptomatic COVID-19 was performed using methodology detailed in Ge 2011[24]. The standardized relative risk reduction (RRR) for pemivibart efficacy versus placebo was calculated after adjusting for age group (≥55 years; <55 years) as a baseline covariate. A stratified Cox proportional-hazards model was used to estimate the hazard ratio (HR) adjusted for the randomization stratification factors. The estimated HR and 95% CI with nominal *P*-value using the score test were derived from the Cox model. Statistical analyses were carried out using SAS (version 9.4 or newer, SAS Institute Inc, Cary, NC). Further details about sample size calculations and statistical analysis plan for cohort B are available as Supplementary Material.

## RESULTS

### Participants

In cohort A, 306 adults with significant immunocompromise were enrolled and received any amount of pemivibart (safety analysis set; **Figure 1**); 298 (97.4%) participants received a full initial dose of pemivibart, comprising the FAS, and 297 (97.1%) participants received a second dose of pemivibart. All participants in cohort A were considered at high risk of COVID-19 disease progression and had immunocompromising conditions, of which taking immunosuppressive medication (66.0%) was the most frequent (**Table 1**). The majority were female (61.1%), White (85.6%), and not Hispanic or Latinx (93.5%). Median age was 59 years, with 31% aged ≥65 years. At baseline, nearly all participants (97.7%) were positive for antibodies to the SARS-CoV-2 S-protein (denoting prior infection or vaccination) and 49.0% were positive for antibodies to N-protein (denoting prior infection). Three asymptomatic participants were confirmed RT-PCR-positive for SARS-CoV-2 at baseline following central laboratory testing.

**Figure 1.**
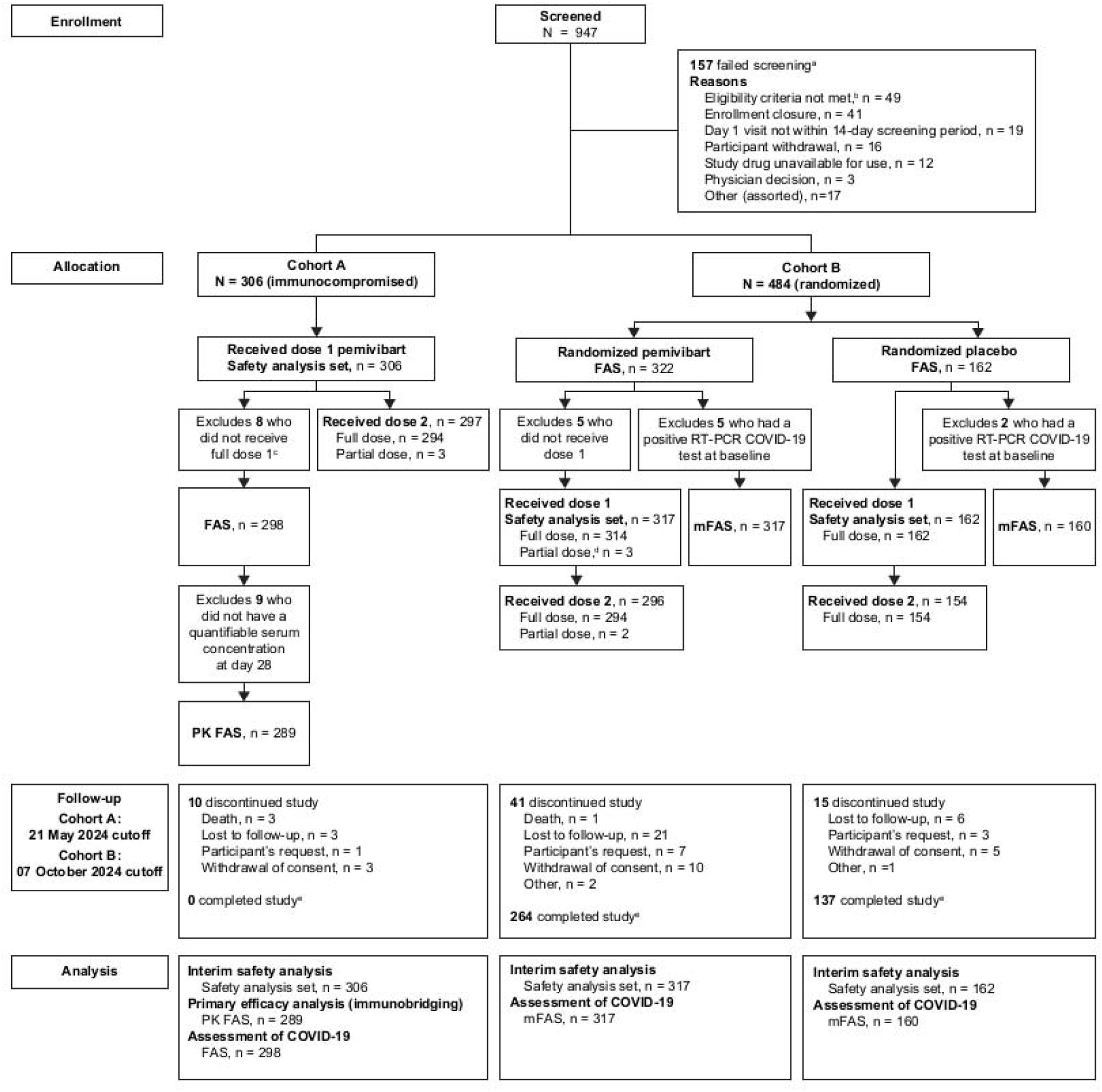
Participant disposition (cohorts A and **B)** Abbreviations: FAS, full analysis set; mFAS, modified full analysis set; PK, pharmacokinetics; RT-PCR, reverse transcription-polymerase chain reaction. ^a^Represents unique events, not individual people. ^b^The most frequent reasons for not meeting the eligibility criteria were no significant immune compromise or no risk of exposure to SARS-CoV-2 identified (n=13); serious concomitant systemic disease or condition that may lead to hospitalization or death, confound study results, or confer additional risk to participant (n=7); and prior known or suspected SARS-CoV-2 infection within 120 days before randomization (n=7). ^c^Reasons for not receiving a full initial dose in cohort A included adverse events (n=5) and issues related to intravenous line placement (n=3). ^d^Reasons for not receiving a full initial dose in cohort B included adverse events (n=2) and issues related to intravenous line placement (n=1). ^e^The study is ongoing through the month 12 visit. Follow-up is shown through 6 months for cohort A and through 12 months for cohort B, as these data were available at the time of this publication.

In cohort B, 484 adults were randomized, and 479 (99.0%) received an initial dose of pemivibart (n=317) or placebo (n=162; safety analysis set; **Figure 1**); 296 and 154 participants received a second dose of pemivibart or placebo, respectively. At the time of the 12-month clinical efficacy analysis, 401 (83%) of participants had completed the study, 56 (12%) had discontinued primarily because of loss of follow-up or withdrawal of consent, and data were unavailable for 27 (6%) participants who had passed the 12-month follow up timepoint as of 07 October 2024. Demographic and baseline characteristics in cohort B were generally well-balanced between treatment arms (**Table 1**). Among all randomized, the majority were female (53.1%), White (63.8%), and not Hispanic or Latinx (69.0%). Median age was 48 years, with 18% aged ≥65 years. Moreover, 64.7% of participants had risk factors for severe/critical COVID-19, the most common of which were obesity (40.1%), age ≥55 years (36.8%), cardiac disease (22.5%), and diabetes (9.1%). Most participants (94.6%) had no history of COVID-19 vaccination or infection (per self-report) within 240 days before randomization; 98.8% and 85.1% were positive for antibodies to the SARS-CoV-2 S-protein and N-protein, respectively, at baseline. Seven asymptomatic participants were confirmed by central laboratory testing as RT-PCR-positive for SARS-CoV-2 at baseline and were excluded from the clinical efficacy analysis (mFAS).

### Safety

#### Cohort A

As of 21 May 2024, TEAEs were reported for 204/306 (66.7%) participants in cohort A; most were classified as mild or moderate severity (**Table 2**). The most frequently reported TEAEs were viral infection (7.8%) and upper respiratory tract infection (7.5%). Serious TEAEs were reported in 35 (11.4%) participants, of which 2 (0.7%) were grade 4 anaphylactic reactions that occurred at redosing and were considered related to pemivibart. In addition, two non-serious infusion-related/hypersensitivity reactions after the first dose, treated with oral diphenhydramine, were reclassified during the regulatory review as anaphylaxis per Sampson’s criteria[25]. More details about the anaphylaxis events are in Supplementary Materials. At the safety cutoff date, there were 3 deaths, including 2 (suicide and unknown cause) that occurred at 92 days after the first dose (no redosing), and 1 (cerebrovascular accident) that occurred at 229 days after the first dose (132 days after the second dose). The most frequently reported study-drug-related TEAEs were IRRs (3.6%). Overall, 25 (8.2%) participants reported symptoms of IRRs and/or HSRs within 24 hours of initial dosing and 12 (4.0%) within 24 hours of redosing (**Table 3**). Seven (2.3%) participants had a study drug-related TEAE resulting in discontinuation of pemivibart (**Table 2; Supplementary Table 1**).

**Table 2.** Summary of Treatment-Emergent Adverse Events (Cohorts A and B, Safety Analysis Set)

| Adverse Event Category, <sup>a</sup> n (%) | Cohort A | Cohort B |  |
| --- | --- | --- | --- |
|  | Pemivibart<br>n=306 | Pemivibart<br>n=317 | Placebo<br>n=162 |
| **Any TEAEs<sup>b</sup>** | 204 (66.7) | 133 (42.0) | 67 (41.4) |
| Grade 1 (mild) | 91 (29.7) | 82 (25.9) | 44 (27.2) |
| Grade 2 (moderate) | 78 (25.5) | 44 (13.9) | 22 (13.6) |
| Grade 3 (severe) | 27 (8.8) | 5 (1.6) | 0 |
| Grade 4 (life threatening) | 5 (1.6) | 1 (0.3) | 1 (0.6) |
| Grade 5 (fatal) | 3 (1.0) | 1 (0.3) | 0 |
| **Most frequent TEAEs (≥5% of participants)** |  |  |  |
| Viral infection | 24 (7.8) | 23 (7.3) | 20 (12.3) |
| Upper respiratory tract infection | 23 (7.5) | 26 (8.2) | 15 (9.3) |
| Influenza-like illness | 13 (4.2) | 17 (5.4) | 9 (5.6) |
| **Serious TEAEs** | 35 (11.4) | 6 (1.9) | 2 (1.2) |
| **TEAEs leading to study treatment interruption<sup>c</sup>** | 20 (6.5) | 5 (1.6) | 0 |
| **TEAEs leading to study treatment discontinuation<sup>d</sup>** | 7 (2.3) | 5 (1.6) | 0 |
| **Any study drug-related TEAEs<sup>b</sup>** | 34 (11.1) | 15 (4.7) | 0 |
| Grade 1 (mild) | 22 (7.2) | 12 (3.8) | 0 |
| Grade 2 (moderate) | 10 (3.3) | 3 (0.9) | 0 |
| Grade 3 (severe) | 0 | 0 | 0 |
| Grade 4 (life threatening) | 2 (0.7) | 0 | 0 |
| Grade 5 (fatal) | 0 | 0 | 0 |
| **Most frequent study drug-related TEAEs ($\geq 1\%$ of participants)** |  |  |  |
| Infusion-related reaction | 11 (3.6) | 7 (2.2) | 0 |
| Fatigue | 5 (1.6) | 0 | 0 |
| Headache | 5 (1.6) | 0 | 0 |
| Infusion-site bruising | 3 (1.0) | 0 | 0 |
| Infusion-site erythema | 3 (1.0) | 0 | 0 |
| **Study drug-related serious TEAE** | 2 (0.7) | 0 | 0 |
| **Study drug-related TEAE leading to study treatment interruption<sup>c</sup>** | 14 (4.6) | 4 (1.3) | 0 |
| **Study drug-related TEAE leading to study treatment discontinuation<sup>d</sup>** | 7 (2.3) | 3 (0.9) | 0 |

Abbreviations: TEAE, treatment emergent adverse event.
<sup>a</sup>Missing relationship assessments were assumed to be 'related'.
<sup>b</sup>Missing severity assessments were not imputed.
<sup>c</sup>Treatment interruption includes any participant who began dosing on day 1, did not complete the dose, but was subsequently redosed; or who had a dose interrupted on day 1 or month 3 but restarted and completed the dose.
<sup>d</sup>Study treatment discontinuation included any participant who began dosing on day 1 but did not finish and was not redosed at month 3, received a full dose on day 1 but was not redosed at month 3, or received a full dose on day 1 and began redosing at month 3 but did not finish. Does not include participants who had a dose interrupted but restarted and completed the dose or had the day 1 dose interrupted/discontinued but was subsequently redosed fully at month 3.

**Table 3.** Summary of Treatment-Emergent Infusion-Related or Hypersensitivity Reactions (IRRs/HSRs) Within 24 Hours of Dosing (Cohorts A and B, Safety Analysis Set and Safety Redosing Set)

| Adverse Event Category | Cohort A | Cohort B |  |
| --- | --- | --- | --- |
|  | Pemivibart | Pemivibart | Placebo |
| **Initial dosing** | **n=306** | **n=317** | **n=162** |
| Any IRRs/HSRs within 24 hours,<sup>a</sup> n (%) | 25 (8.2) | 4 (1.3) | 1 (0.6) |
| Grade 1 (mild) | 17 (5.6) | 3 (0.9) | 1 (0.6) |
| Grade 2 (moderate) | 8 (2.6) | 1 (0.3) | 0 |
| **Redosing<sup>b</sup>** | **n=297** | **n=296** | **n=154** |
| Any IRRs/HSRs within 24 hours,<sup>a</sup> n (%) | 12 (4.0) | 8 (2.7) | 0 |
| Grade 1 (mild) | 6 (2.0) | 7 (2.4) | 0 |
| Grade 2 (moderate) | 4 (1.3) | 1 (0.3) | 0 |
| Grade 4 (life threatening) | 2 (0.7) | 0 | 0 |

<sup>a</sup>IRRs/HSRs include altered mental status, anaphylactic reaction, arrhythmia, atrial fibrillation, bradycardia, brain fog, chest pain, chills, dermatitis, diaphoresis, diarrhea, dizziness, fatigue, fever, headache, hypersensitivity, hypertension, infusion-related hypersensitivity reaction, infusion-related reaction, mental status changes, myalgia, nausea, paresthesia, presyncope, sinus tachycardia, syncope, tachycardia, throat irritation, tremor, vasovagal reaction, weakness.
<sup>b</sup>Includes all participants who received two doses (any amount of study drug at both initial dosing and redosing).

#### Cohort B

As of 21 May 2024, in cohort B, the incidence of TEAEs was 42.0% (133/317) in the pemivibart group and 41.4% (67/162) in the placebo group (**Table 2**). Like cohort A, most TEAEs were classified as mild or moderate severity; the most frequent TEAEs were viral infection (pemivibart, 7.3%; placebo, 12.3%) and upper respiratory tract infection (pemivibart, 8.2%; placebo, 9.3%). Serious TEAEs were reported in 6 (1.9%) of the pemivibart group and 2 (1.2%) of the placebo group; none were considered related to study drug. There was 1 death due to congestive cardiac failure in the pemivibart group that occurred 183 days after the first dose (no redosing). In the pemivibart group, study-drug-related TEAEs were reported in 15 (4.7%) participants, including 7 (2.2%) IRRs, and 3 (0.9%) that led to treatment discontinuation (**Supplementary Table 1**). Overall, 5 participants reported symptoms of IRRs/HSRs within 24 hours of initial dosing (pemivibart, n=4 [1.3%]; placebo, n=1 [0.6%]) and 8 within 24 hours of redosing (all in the pemivibart group, n=8 [2.7%]; **Table 3**); no severe or serious IRRs/HSRs or anaphylaxis cases were reported.

### Immunobridging

For cohort A, the efficacy of pemivibart was assessed through immunobridging to historical data from the EVADE study, which provided evidence of clinical efficacy of adintrevimab, the parent mAb of pemivibart. The results showed that the geometric mean ratio between the calculated sVNA titer for pemivibart against the JN.1 variant at day 28 (based on a pseudovirus neutralization assay [IC_50_] value of 74.6 ng/ml) and the calculated titer for adintrevimab against Delta (based on an authentic virus neutralization assay IC_50_ value of 7 ng/mL) was 0.70 (90% CI, 0.68-0.72)[20]. A supplementary immunobridging analysis demonstrated that pemivibart titers were consistent with the titer levels associated with efficacy in prior clinical trials evaluating certain mAbs for the prevention of COVID-19[20].

### COVID-19 Assessments

#### Cohort A

In cohort A (FAS), the composite incidence of RT-PCR-confirmed symptomatic COVID-19, COVID-related hospitalization, or all-cause mortality was 3/298 (1.0%; no deaths) through month 3 and 11/298 (3.7%; 2 deaths) through month 6 (**Table 4**). There were no COVID-19-related hospitalizations through month 6. The time-to-event analysis (**Figure 2**) showed that the estimated probability of the composite endpoint was 0.8% (95% CI, 0.2-2.7) through day 90 and 4.1% (95% CI, 2.1-7.1) through month 6.

**Table 4.**
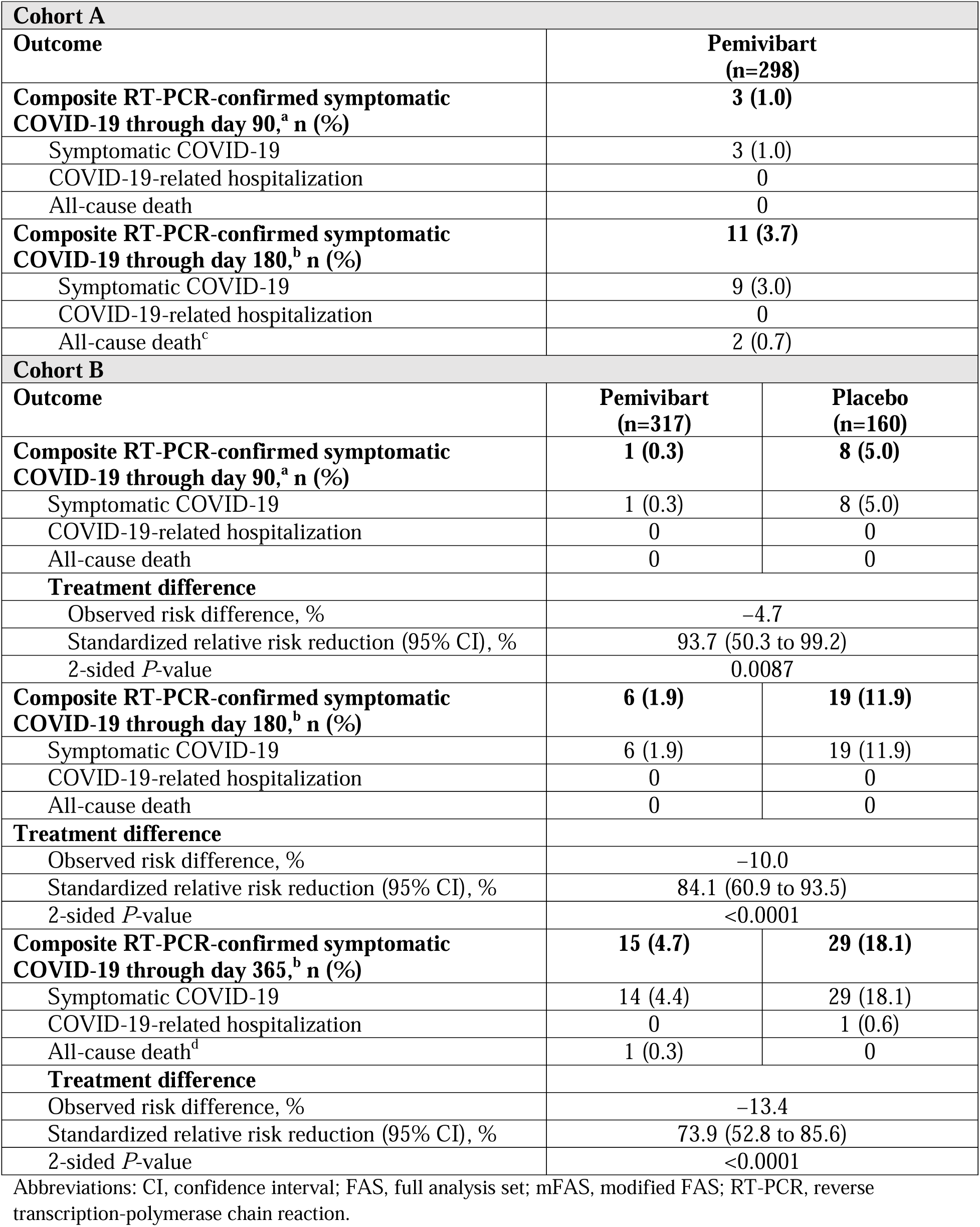
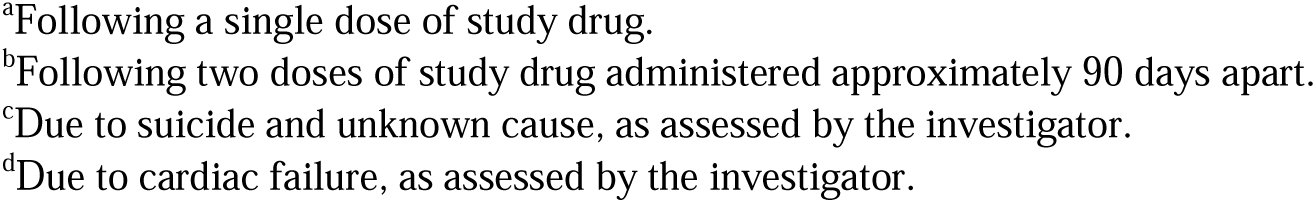
Composite RT-PCR-Confirmed Symptomatic COVID-19 (Cohort A, FAS; Cohort B, mFAS)

| Cohort A |  |  |
| --- | --- | --- |
| Outcome | Pemivibart<br>(n=298) |  |
| Composite RT-PCR-confirmed symptomatic COVID-19 through day 90,<sup>a</sup> n (%) | 3 (1.0) |  |
| Symptomatic COVID-19 | 3 (1.0) |  |
| COVID-19-related hospitalization | 0 |  |
| All-cause death | 0 |  |
| Composite RT-PCR-confirmed symptomatic COVID-19 through day 180,<sup>b</sup> n (%) | 11 (3.7) |  |
| Symptomatic COVID-19 | 9 (3.0) |  |
| COVID-19-related hospitalization | 0 |  |
| All-cause death<sup>c</sup> | 2 (0.7) |  |
| Cohort B |  |  |
| Outcome | Pemivibart<br>(n=317) | Placebo<br>(n=160) |
| Composite RT-PCR-confirmed symptomatic COVID-19 through day 90,<sup>a</sup> n (%) | 1 (0.3) | 8 (5.0) |
| Symptomatic COVID-19 | 1 (0.3) | 8 (5.0) |
| COVID-19-related hospitalization | 0 | 0 |
| All-cause death | 0 | 0 |
| Treatment difference |  |  |
| Observed risk difference, % | −4.7 |  |
| Standardized relative risk reduction (95% CI), % | 93.7 (50.3 to 99.2) |  |
| 2-sided *P*-value | 0.0087 |  |
| Composite RT-PCR-confirmed symptomatic COVID-19 through day 180,<sup>b</sup> n (%) | 6 (1.9) | 19 (11.9) |
| Symptomatic COVID-19 | 6 (1.9) | 19 (11.9) |
| COVID-19-related hospitalization | 0 | 0 |
| All-cause death | 0 | 0 |
| Treatment difference |  |  |
| Observed risk difference, % | −10.0 |  |
| Standardized relative risk reduction (95% CI), % | 84.1 (60.9 to 93.5) |  |
| 2-sided *P*-value | <0.0001 |  |
| Composite RT-PCR-confirmed symptomatic COVID-19 through day 365,<sup>b</sup> n (%) | 15 (4.7) | 29 (18.1) |
| Symptomatic COVID-19 | 14 (4.4) | 29 (18.1) |
| COVID-19-related hospitalization | 0 | 1 (0.6) |
| All-cause death<sup>d</sup> | 1 (0.3) | 0 |
| Treatment difference |  |  |
| Observed risk difference, % | −13.4 |  |
| Standardized relative risk reduction (95% CI), % | 73.9 (52.8 to 85.6) |  |
| 2-sided *P*-value | <0.0001 |  |

Abbreviations: CI, confidence interval; FAS, full analysis set; mFAS, modified FAS; RT-PCR, reverse transcription-polymerase chain reaction.
<sup>a</sup>Following a single dose of study drug.
<sup>b</sup>Following two doses of study drug administered approximately 90 days apart.
<sup>c</sup>Due to suicide and unknown cause, as assessed by the investigator.
<sup>d</sup>Due to cardiac failure, as assessed by the investigator.

**Figure 2.**
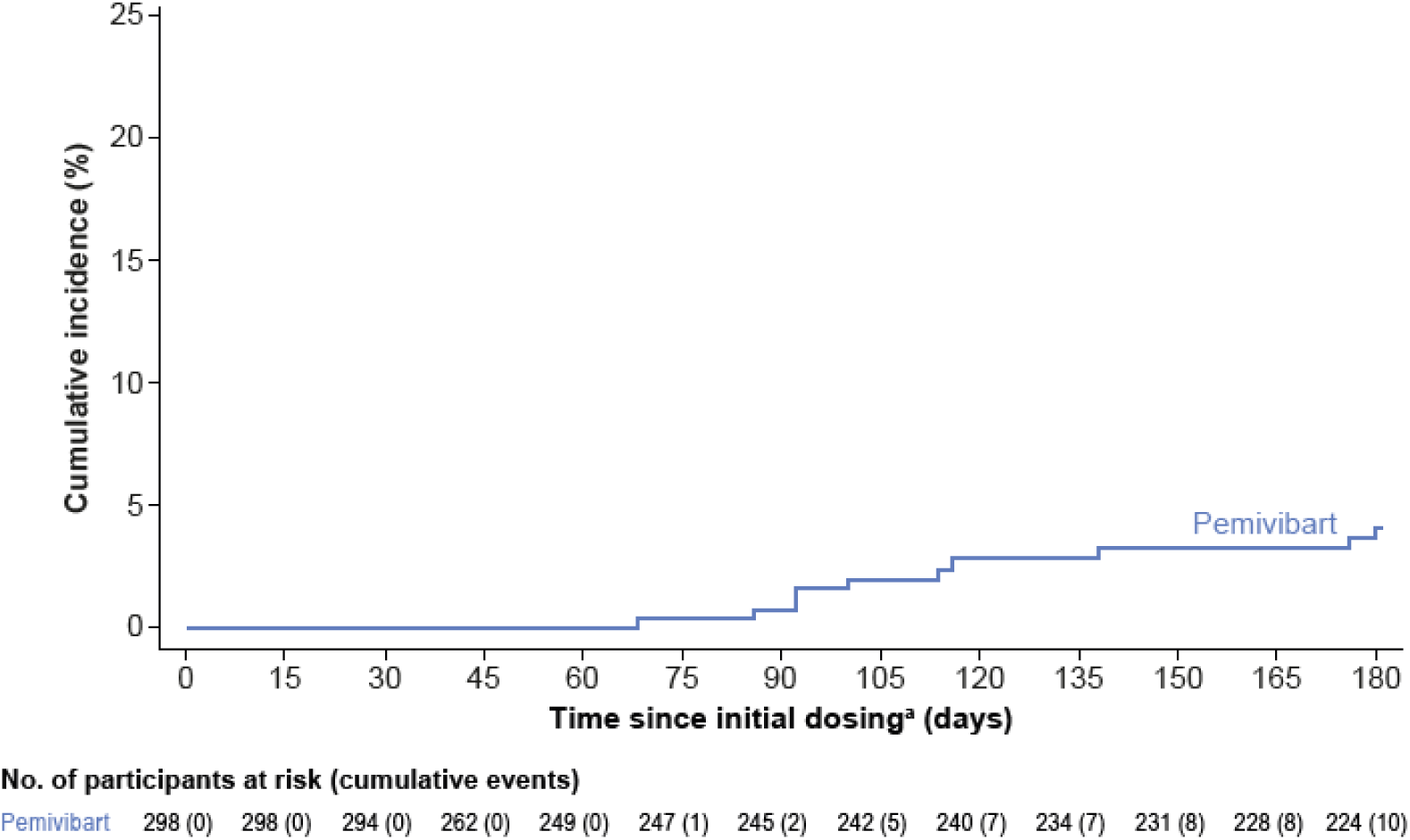
Cumulative incidence of RT-PCR-confirmed COVID-19, COVID-related hospitalization, and all-cause mortality through day 180, cohort A, FAS. Abbreviations: FAS, full analysis set; RT-PCR, reverse transcription-polymerase chain reaction. Time to RT-PCR-confirmed symptomatic COVID-19 is calculated as date of event/censoring – date of randomization + 1. Participants who did not have the defined event on or before the above censoring date are censored at the earliest of the end-of-study date, 180-day follow-up, date of participants receipt of COVID-19 vaccination, and analysis cutoff date. One participant in cohort A who received postdose COVID-19 vaccination was censored at the time of COVID-19 vaccination. ^a^A second dose of study drug was administered approximately 90 days after initial dosing.

#### Cohort B

In cohort B (mFAS), the composite incidence of RT-PCR-confirmed symptomatic COVID-19, COVID-related hospitalization, and all-cause mortality through month 3 was 1/317 (0.3%) in the pemivibart group and 8/160 (5.0%) in the placebo group, representing a 93.7% RRR (95% CI, 50.3-99.2; nominal *P*=.0087) with pemivibart (**Table 4**). Through month 6, 6 (1.9%) participants in the pemivibart group and 19 (11.9%) in the placebo group met the endpoint, representing an 84.1% RRR (95% CI, 60.9-93.5; nominal *P*<.0001) with pemivibart. There were no COVID-19-related hospitalizations or all-cause deaths through month 6. Through month 12, 15 (4.7%, 1 death due to cardiac failure) participants in the pemivibart group and 29 (18.1%, 1 COVID-19 related hospitalization) in the placebo group met the composite endpoint, representing a 73.9% RRR (95% CI, 52.8-85.6; nominal *P*<.0001). Importantly, these risk reductions span through JN.1 and the most recent dominant variant KP.3.1.1. In the time-to-event analysis (**Figure 3**), the hazard ratio (HR) of the composite endpoint through month 12 after 2 doses of pemivibart was 0.25 (95% CI, 0.13-0.47; nominal *P*=<.0001) versus placebo.

**Figure 3.**
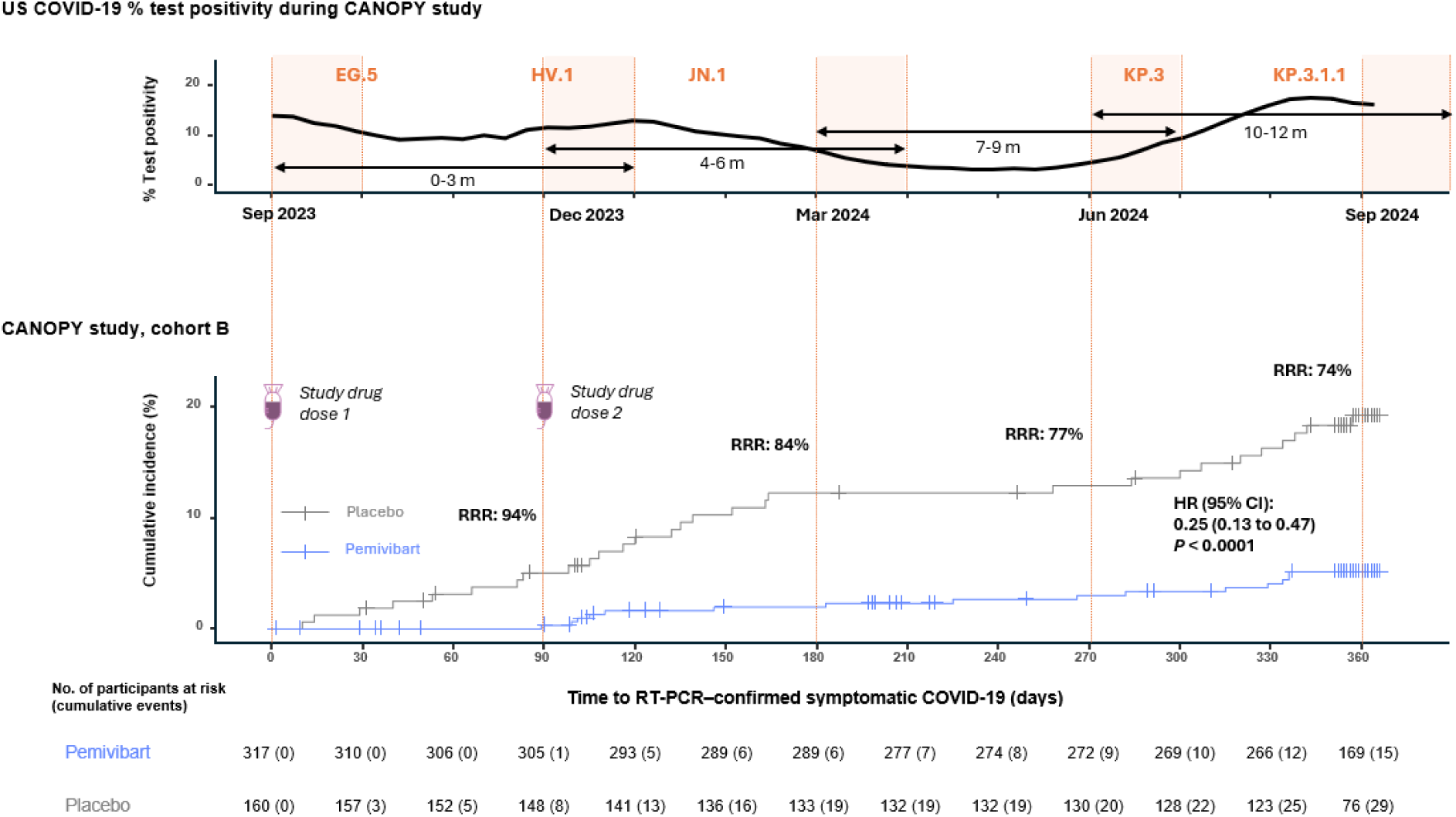
Cumulative incidence of RT-PCR-confirmed COVID-19, COVID-related hospitalization, and all-cause mortality through day 180, cohort B, mFAS, shown through percent COVID-19 test positivity in the United States during the CANOPY study Abbreviations: mFAS, modified full analysis set; RT-PCR, reverse transcription-polymerase chain reaction; RRR, relative risk reduction. Shaded boxes in the top graph represent the overlap of time periods of cohorts A-B in the variant landscape. Time to RT-PCR-confirmed symptomatic COVID-19 was calculated as date of event/censoring – date of randomization + 1. Participants who did not have the defined event on or before the above censoring date were censored at the earliest of the end-of-study date, 365-day follow-up, and analysis cutoff date. The standardized relative risk reduction for pemivibart efficacy versus placebo was calculated for cumulative period through 90, 180, 270 and 365 days, respectively, adjusting for age group (≥55 years; <55 years) as a baseline covariate, using methodology detailed in Ge 2011[24]. A second dose of study drug was administered approximately 90 days after initial dosing.

### Pharmacokinetics and Immunogenicity

In the pemivibart population PK (popPK) model, a linear, 2-compartment model with zero-order IV input and allometric scaling of clearance and volume of the central compartment provided a robust fit to the pooled data with reliable estimation of popPK parameters for the CANOPY study (**Table 5**). The estimated median terminal elimination half-life of pemivibart was 49 days at the latest interim analysis. Treatment-emergent ADAs were detected in 6 (2%) participants in cohort A and 3 (1%) in cohort B at various post-baseline timepoints, all with very low titers (≤minimum required dilution). See **Supplementary Materials** for PopPK covariate assessment results and details on ADAs.

**Table 5.**
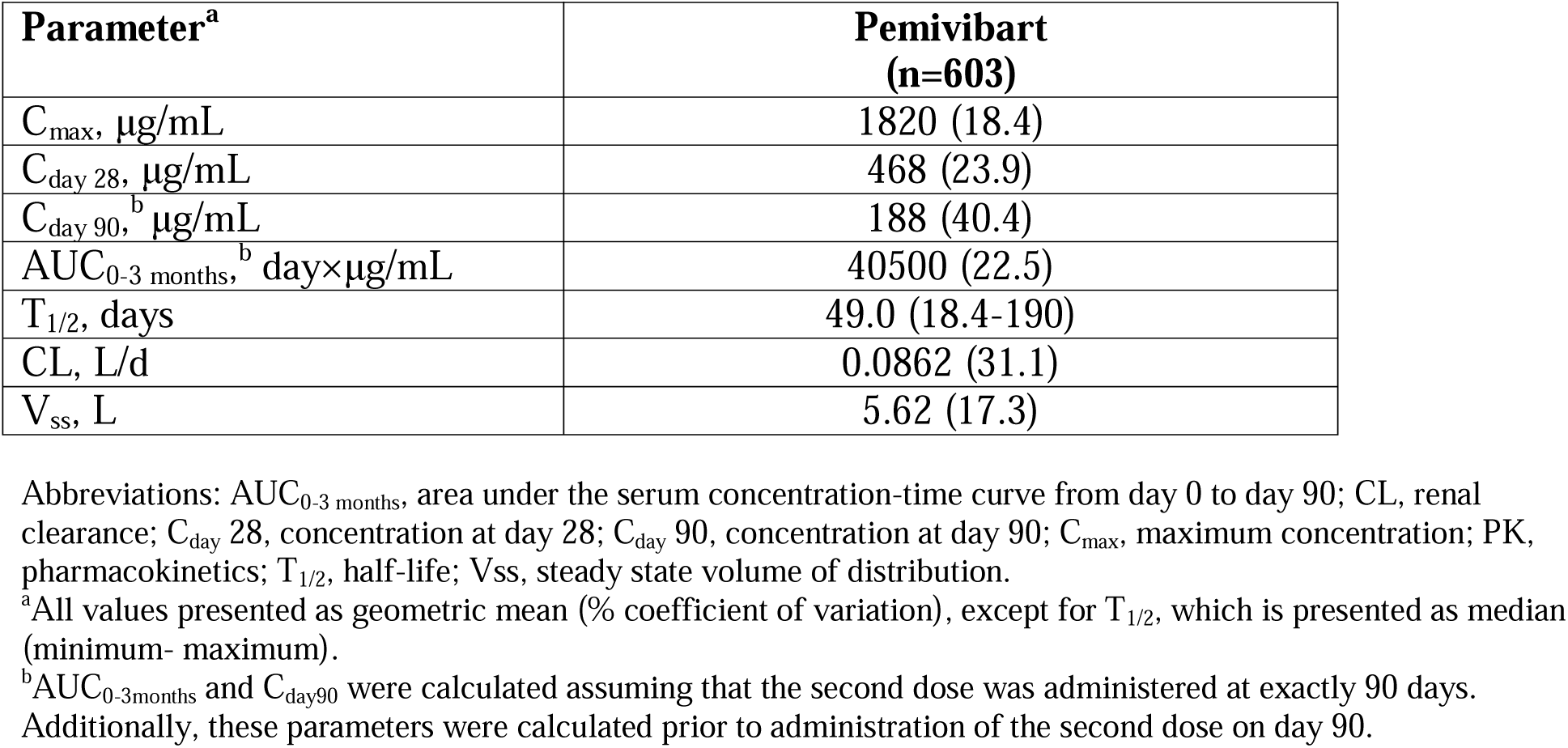
Summary Statistics of PK Parameters Following a Single 4500 mg IV Dose of Pemivibart (Pooled Cohorts A and B, Population PK Analysis)

## DISCUSSION

Pemivibart is the first mAb to receive EUA for prevention of COVID-19 in certain immunocompromised people based on a rapid immunobridging trial design[20]. The primary analysis in the cohort of immunocompromised participants used calculated sVNA titers as a surrogate for clinical efficacy[20]. Here we add to the totality of evidence showing that 2 IV infusions of pemivibart administered 90 days apart were well-tolerated by most participants, and the composite incidence of RT-PCR-confirmed symptomatic COVID-19, COVID-related hospitalization, or all-cause mortality was lower through 12 months spanning contemporary variants in the pemivibart group versus the placebo group in participants without immunocompromise.

Clinical trials of mAbs as prophylaxis for COVID-19 have largely been conducted in non-immunocompromised individuals[12–14]. Therefore, a primary objective of CANOPY was to evaluate safety and tolerability of pemivibart in a large cohort of immunocompromised individuals. Over a 6-month follow-up period, pemivibart was generally well-tolerated. The incidence of TEAEs was similar in the pemivibart and placebo groups in cohort B, and most TEAEs in both cohorts were classified as mild or moderate in severity. IRRs/HSRs, including anaphylaxis, are known risks of COVID-19 mAbs[26]. We noted that a larger proportion of immunocompromised (8.2%) than immunocompetent participants (1.3%) reported IRRs/HSRs at the initial dosing of pemivibart. Four participants (all in the immunocompromised cohort) experienced anaphylactic reactions, of which 2 were serious and occurred within 24 hours of the second dose. All other IRRs/HSRs were mild or moderate. The differences between cohorts may be due to the underlying immunocompromise, which can alter both innate and adaptive immune responses[8]. The incidence of IRRs/HSRs in CANOPY was consistent with the range (<0.1% to 13%) observed in non-immunocompromised participants who received IV mAbs in previous clinical trials for the prevention or treatment of COVID-19[12,27–30]. IRRs/HSRs rates could also be partly due to the infusion duration, as initially most participants were dosed and redosed with a 30-minute infusion. Following safety data review, a 60-minute infusion was recommended for the remaining participants awaiting redosing.

CANOPY exploratory clinical efficacy data are encouraging. Nearly all participants (>98%) had a history of COVID-19 infection or vaccination. Yet despite that, 2 doses of pemivibart administered 90 days apart provided additional protection against symptomatic COVID-19 caused by contemporary variants in this highly vaccinated population. The composite incidence of RT-PCR-confirmed symptomatic COVID-19, COVID-related hospitalization, or all-cause mortality through month 6 was low at 3.7% in immunocompromised participants, and 1.9% in non-immunocompromised participants who received pemivibart compared with 11.9% in non-immunocompromised participants who received placebo. In a real-world observational study, immunocompromised individuals accounted for >20% of COVID-19 hospitalizations and deaths, although they represented only 3.9% of the study population[3]. In CANOPY, a diverse immunocompromised population was enrolled, with 66% of people taking immunosuppressive medications, 13% having hematologic malignancies, 12% having primary immunodeficiency, and 11% having received SOT. Importantly, there were no COVID-related hospitalizations in these participants, suggesting that pemivibart may provide protection for those who have a higher risk of severe outcomes[31].

In cohort B, the 74% RRR with pemivibart over 12 months is clinically significant, particularly given the shorter duration of efficacy evaluated in other clinical trials of mAbs in a non-immunocompromised population, including adintrevimab (71.0%)[17], tixagevimab/cilgavimab (76.7%)[13], and casirivimab/imdevimab (81.4%)[14]. The time-to-event analysis showed an increase in RT-PCR-confirmed symptomatic COVID-19 cases in both cohorts, which coincided with surges in COVID-19 cases globally and in the United States in December 2023-January 2024 and July-August 2024 [1,2].

Overall, these data underscore the ongoing need for preventive measures, especially during periods of high exposure to SARS-CoV-2. Future mAbs will likely depend on immunobridging analyses to stay relevant in the face of rapidly evolving SARS-CoV-2 in an endemic era in which hospitalization can no longer be relied upon as an endpoint for a prevention study. The clinical results from CANOPY support the usefulness of the immunobridging approach used for the pemivibart EUA and may indicate further analysis to investigate a lower titer threshold for protection against symptomatic COVID-19 is needed.

Our study has limitations. Cohort A was open label for ethical reasons because of the population’s high-risk status. Altogether, safety and efficacy of pemivibart were evaluated in 306 individuals with significant immunocompromise; however, this may not be wholly representative of the immunocompromised population, especially for those with low to no enrollment such as Hispanic and Latinx people and people with B-cell depletion, chimeric antigen receptor T-cell therapy, or hematopoietic stem cell transplant. Black or African American and Hispanic and Latinx populations were well represented in cohort B. Finally, as for all COVID-19 clinical trials, the participants were primarily exposed to the variant(s) circulating at the time of the study; therefore, efficacy results may not be generalizable to emerging variants. Continued post-marketing surveillance and monitoring of pemivibart activity against new circulating variants is ongoing[32].

These data from CANOPY support pemivibart as a preventive option in an evolving variant landscape against COVID-19 for individuals with significant immunocompromise, such as SOT recipients, those with hematological malignancies, and those taking immunosuppressive medications, who continue to have both a potential suboptimal response to vaccines and a higher risk for severe COVID-19 outcomes.

## Notes

## Author contributions

M.P., A.H., Y.L., I.Y., E.C., and K.N. contributed to study design. M.P., A.H., Y.L., I.Y., D.G., K.N., E.C., and A.P. were involved in protocol development. J.C. was a principal investigator. M.P., A.H., K.M., and N.B. were responsible for medical monitoring. All authors contributed to data interpretation and were involved in drafting and critically revising the manuscript, and all authors approved the final version and are accountable for the accuracy and integrity of the manuscript. All authors had full access to the data in the study and had final responsibility for the decision to submit for publication. Y. L., D.G., and K.N. verified the data.

## Acknowledgments

The authors thank the investigators, site personnel, and the trial participants and their families. We thank the independent data monitoring committee for their review of safety data. Writing assistance for the manuscript was provided by Georgiana Manica, PhD, and Jean Turner of Parexel, and was funded by Invivyd, Inc. David A. Wilfret, MD, of Duke University Medical Center, contributed to the editorial review of the paper.

## Data availability

As this trial is ongoing, data are not publicly available.

## REFERENCES

1. World Health Organization. WHO COVID-19 dashboard. Available at: https://data.who.int/dashboards/covid19/cases?n=c. Accessed 23 September 2024.

2. Centers for Disease Control and Prevention. CDC COVID-19 data tracker. Available at: https://covid.cdc.gov/covid-data-tracker/#hospitalizations-landing. Accessed September 23, 2024.

3. Evans RA, Dube S, Lu Y, et al. Impact of COVID-19 on immunocompromised populations during the Omicron era: insights from the observational population-based INFORM study. Lancet Reg Health Eur 2023; 35.

4. Ketkar A, Willey V, Pollack M, et al. Assessing the risk and costs of COVID-19 in immunocompromised populations in a large United States commercial insurance health plan: the EPOCH-US Study. Curr Med Res Opin 2023; 39: 1103–18.

5. Kim L, Garg S, O’Halloran A, et al. Risk factors for intensive care unit admission and in-hospital mortality among hospitalized adults identified through the US Coronavirus Disease 2019 (COVID-19)-Associated Hospitalization Surveillance Network (COVID-NET). Clin Infect Dis 2021; 72: e206–e14.

6. Singson JRC, Kirley PD, Pham H, et al. Factors associated with severe outcomes among immunocompromised adults hospitalized for COVID-19 - COVID-NET, 10 States, March 2020-February 2022. MMWR Morb Mortal Wkly Rep 2022; 71: 878–84.

7. DeCuir J, Payne AB, Self WH, et al. Interim effectiveness of updated 2023-2024 (monovalent XBB.1.5) COVID-19 vaccines against COVID-19-associated emergency department and urgent care encounters and hospitalization among immunocompetent adults aged ≥18 years - VISION and IVY Networks, September 2023-January 2024. MMWR Morb Mortal Wkly Rep 2024; 73: 180–8.

8. DeWolf S, Laracy JC, Perales MA, Kamboj M, van den Brink MRM, Vardhana S. SARS-CoV-2 in immunocompromised individuals. Immunity 2022; 55: 1779–98.

9. Link-Gelles R, Rowley EAK, DeSilva MB, et al. Interim effectiveness of updated 2023-2024 (monovalent XBB.1.5) COVID-19 vaccines against COVID-19-associated hospitalization among adults aged ≥18 years with immunocompromising conditions - VISION Network, September 2023-February 2024. MMWR Morb Mortal Wkly Rep 2024; 73: 271–6.

10. Corti D, Purcell LA, Snell G, Veesler D. Tackling COVID-19 with neutralizing monoclonal antibodies. Cell 2021; 184: 3086–108.

11. Shoham S, Batista C, Ben Amor Y, et al. Vaccines and therapeutics for immunocompromised patients with COVID-19. EClinicalMedicine 2023; 59: 101965.

12. Cohen MS, Nirula A, Mulligan MJ, et al. Effect of Bamlanivimab vs Placebo on Incidence of COVID-19 Among Residents and Staff of Skilled Nursing and Assisted Living Facilities: A Randomized Clinical Trial. Jama 2021; 326: 46–55.

13. Levin MJ, Ustianowski A, De Wit S, et al. Intramuscular AZD7442 (tixagevimab-cilgavimab) for prevention of Covid-19. N Engl J Med 2022; 386: 2188–200.

14. O’Brien MP, Forleo-Neto E, Musser BJ, et al. Subcutaneous REGEN-COV antibody combination to prevent Covid-19. N Engl J Med 2021; 385: 1184–95.

15. Planas D, Saunders N, Maes P, et al. Considerable escape of SARS-CoV-2 Omicron to antibody neutralization. Nature 2022; 602: 671–5.

16. Ison MG, Popejoy M, Evgeniev N, et al. Efficacy and safety of adintrevimab (ADG20) for the treatment of high-risk ambulatory patients with mild or moderate coronavirus disease 2019: results from a phase 2/3, randomized, placebo-controlled trial (STAMP) conducted during Delta predominance and early emergence of Omicron. Open Forum Infect Dis 2023; 10: ofad279.

17. Ison MG, Weinstein DF, Dobryanska M, et al. Prevention of COVID-19 following a single intramuscular administration of adintrevimab: results from a phase 2/3 randomized, double-blind, placebo-controlled trial (EVADE). Open Forum Infect Dis 2023; 10: ofad314.

18. Rappazzo CG, Tse LV, Kaku CI, et al. Broad and potent activity against SARS-like viruses by an engineered human monoclonal antibody. Science 2021; 371: 823–9.

19. Wec AZ, Wrapp D, Herbert AS, et al. Broad neutralization of SARS-related viruses by human monoclonal antibodies. Science 2020; 369: 731–6.

20. Invivyd, Inc. Fact sheet for healthcare providers: emergency use authorization of pemgarda (pemivibart). Available at: https://www.fda.gov/media/177067/download. Accessed 11 November 2024.

21. Mahoney K, Gupta D, Li Y, et al. Preliminary safety results from a phase 1 first in human study of VYD222: an extended half-life monoclonal antibody (mAb) in development for COVID-19 prevention. Presented at: ID Week 2023; 11–13 October 2023; Boston, MA.

22. FDA. Emergency Use Authorization (EUA) for PEMGARDA Center for Drug Evaluation and Research (CDER) Review Memorandum. Available at: https://www.fda.gov/media/181308/download#:∼:text=PEMGARDA%20received%20Emergency%20Use. Accessed October 07, 2024.

23. ClinicalTrials.gov. A study to investigate the prevention of COVID-19 withVYD222 in adults with immune compromise and in participants aged 12 years or older who are at risk of exposure to SARS-CoV-2. Available at: https://classic.clinicaltrials.gov/ct2/show/NCT06039449. Accessed 23 September 2024.

24. Ge M, Durham LK, Meyer RD, Xie W, Thomas N. Covariate-adjusted difference in proportions from clinical trials using logistic regression and weighted risk differences. Drug Inf J 2011; 45: 481–93.

25. Sampson HA, Muñoz-Furlong A, Campbell RL, et al. Second symposium on the definition and management of anaphylaxis: summary report--Second National Institute of Allergy and Infectious Disease/Food Allergy and Anaphylaxis Network symposium. J Allergy Clin Immunol 2006; 117: 391–7.

26. Chary M, Barbuto AF, Izadmehr S, Tarsillo M, Fleischer E, Burns MM. COVID-19 therapeutics: use, mechanism of action, and toxicity (vaccines, monoclonal antibodies, and immunotherapeutics). J Med Toxicol 2023; 19: 205–18.

27. Dougan M, Azizad M, Chen P, et al. Bebtelovimab, alone or together with bamlanivimab and etesevimab, as a broadly neutralizing monoclonal antibody treatment for mild to moderate, ambulatory COVID-19. medRxiv 2022: 2022.03.10.22272100.

28. Group A-TfIwC-S. Efficacy and safety of two neutralising monoclonal antibody therapies, sotrovimab and BRII-196 plus BRII-198, for adults hospitalised with COVID-19 (TICO): a randomised controlled trial. Lancet Infect Dis 2022; 22: 622–35.

29. Rosas IO, Bräu N, Waters M, et al. Tocilizumab in Hospitalized Patients with Severe Covid-19 Pneumonia. N Engl J Med 2021; 384: 1503–16.

30. Weinreich DM, Sivapalasingam S, Norton T, et al. REGEN-COV Antibody Combination and Outcomes in Outpatients with Covid-19. N Engl J Med 2021; 385: e81.

31. Razonable RR. Protecting the vulnerable: addressing the COVID-19 care needs of people with compromised immunity. Front Immunol 2024; 15: 1397040.

32. Invivyd, Inc. Invivyd provides detailed virology data and analysis of SARS-CoV-2 structural biology predicting anticipated neutralization activity for PEMGARDA™ (pemivibart). Available at: https://investors.invivyd.com/news-releases/news-release-details/invivyd-provides-detailed-virology-data-and-analysis-sars-cov-2. Accessed 24 September 2024.

