## Supplemental Material for "Safety and Efficacy of Pemivibart, a Long-Acting Monoclonal Antibody, for Prevention of Symptomatic COVID-19: Interim Results From the CANOPY Clinical Trial"

**Table of Contents**


LIST OF TRIAL INVESTIGATORS ... 2
KEY PROTOCOL AMENDMENTS ... 3
SUPPLEMENTARY METHODS ... 5
PARTICIPANTS ... 5
Enrollment ... 5
Inclusion Criteria ... 5
Exclusion Criteria ... 7
PHARMACOKINETICS ... 10
IMMUNOGENICITY ... 10
SAMPLE SIZE ... 10
SUPPLEMENTARY RESULTS ... 12
ANAPHYLAXIS MANAGEMENT ... 12
PopPK MODEL COVARIATE ASSESSMENTS ... 12
IMMUNOGENICITY ... 13
SUPPLEMENTARY REFERENCES ... 14

### LIST OF TRIAL INVESTIGATORS

| Name of Principal Investigator | Institution | City | State |
|---|---|---|---|
| Kevin Oei | Ascada Research | Fullerton | California |
| Codey Bell | Tekton Research | Beaumont | Texas |
| Robert Carter | Coastal Heritage Clinical Research LLC | Hinesville | Georgia |
| Vasundhara Cheekati | Agile Clinical Research Trials, LLC | Atlanta | Georgia |
| Jonathan Cohen | Jadestone Clinical Research | Silver Spring | Maryland |
| Roxana Stoici | GCP, Global Clinical Professionals Elias Research Associates | St. Petersburg | Florida |
| Barry Heller | Long Beach Clinical Trials, LLC | Long Beach | California |
| Gary Kleeman | Skylight Health Research | Burlington | Massachusetts |
| Stacey Layle | Artemis Institute for Clinical Research - San Diego | San Diego | California |
| James Lilly | Zenos Clinical Research | Dallas | Texas |
| Robert Lockwood | Tekton Research | Yukon | Oklahoma |
| Sana Meah | Alliance for Multispecialty Research (AMR) LLC | Oak Brook | Illinois |
| Rakesh Patel | Onsite Clinical Solutions LLC | Salisbury | North Carolina |
| Kyle Rickner | Tekton Research | Edmond | Oklahoma |
| Lawrence Sher | Peninsula Research Associates, Inc | Rolling Hills Estates | California |
| Miguel Trevino | Innovative Research of West Florida, Inc. | Clearwater | Florida |
| Juan Velazquez | Nuovida Research Center Corp | Miami | Florida |
| Debra Weinstein | Science37, Inc. | Morrisville | North Carolina |

### KEY PROTOCOL AMENDMENTS

The protocol was amended 3 times.

**Version 1 (22 June 2023)**

- Original protocol
- No participants were enrolled under this version of the protocol

**Version 2 (7 August 2023)**

- Updated to change dose regimen from a single dose of pemivibart 2500 mg intravenous (IV) to pemivibart 4500 mg IV on day 1 followed by redosing approximately 90 days after day 1
- All participants were enrolled under this version of the protocol

**Version 3 (29 November 2023)**

- Updated cohort B statistical analyses and endpoints: adapted cohort B primary and key secondary endpoints and analyses for reverse transcription-polymerase chain reaction (RT-PCR)-confirmed symptomatic COVID-19
- No updates impacting study participants or study conduct at clinical sites

**Version 4 (15 April 2024)**

- Adapted cohort B RT-PCR-confirmed symptomatic COVID-19 endpoints to be exploratory due to early sponsor unblinding of clinical efficacy data for EUA application prior to the intended timing of analysis
- Clarified cohort A immunobridging hypothesis statement

- Incorporated revisions previously released in the protocol clarification letters, including updates to inclusion criteria 4avii, study drug discontinuation for infusion-related reactions, and COVID-like illness visits
- No updates impacting study participants or study conduct at clinical sites.

### SUPPLEMENTARY METHODS

### PARTICIPANTS

*Enrollment*

The independent data monitoring committee reviewed the safety data from the first 100 participants exposed to pemivibart combined across both cohorts and provided a recommendation for opening cohort B to enrollment of adolescent and pregnant/breastfeeding individuals. However, the sample size target was met before this time, and no adolescents or pregnant/breastfeeding participants were enrolled.

*Inclusion Criteria*

Participants are eligible to be included in the study only if all of the following criteria apply:

1. Is an adult aged ≥18 years or an adolescent aged 12 to <18 years and weighs at least 40 kg at the time of screening
   ***Note***: Adolescent enrollment is allowed only in cohort B, open only upon sponsor communication to sites after review of safety lead-in data and only if permitted by the local health authority.
2. Tests negative for current SARS-CoV-2 infection by local antigen test or RT-PCR at the time of screening
3. For cohort A, meets the criteria for one of the following subgroups:
   a. Seronegative or low titer: Had an inadequate response to a COVID-19 bivalent vaccine booster, as demonstrated by SARS-CoV-2 S antibodies <205 BAU/mL measured within 28 to 60 days after the bivalent booster dose and has not received any subsequent vaccine dose prior to day 1, **OR**
   b. Moderate titer: Had SARS-CoV-2 S antibodies ≥205 to <2500 BAU/mL measured within 28 to 60 days after a COVID-19 bivalent vaccine booster and has not received any subsequent vaccine dose prior to day 1, **OR**
   c. High titer: Had SARS-CoV-2 S antibodies ≥2500 BAU/mL measured within 28 to 60 days after a COVID-19 bivalent vaccine booster dose, has not received any subsequent vaccine dose prior to day 1, and >120 days have elapsed since the last dose, **OR**

d. Unknown titer: Has not received any SARS-CoV-2 vaccine in the 120 days prior to study enrollment and has either not received a COVID-19 bivalent vaccine booster dose or has received a booster but did not have S antibody titer determined within 28 to 60 days post dose, **OR**

e. All comer: Meets other criteria for inclusion in cohort A but does not fulfill criteria 3a, 3b, 3c, or 3d and has not had any SARS-CoV-2 vaccine within 14 days prior to day 1

***Note***: Enrollment of subgroups may be opened or closed upon sponsor communication. SARS-CoV-2 serology testing may be performed at a local lab to determine participant subgroup eligibility

4. Has significant immune compromise OR is at risk of exposure to SARS-CoV-2, as assessed by the investigator, as follows:
   a. For cohort A, has significant immune compromise, defined as any of the following:
      i. Actively treated for solid tumor or hematologic malignancies
      ii. Acute leukemia, chronic lymphocytic leukemia, non-Hodgkin lymphoma, or multiple myeloma (regardless of treatment)
      iii. Solid organ transplant recipient taking immunosuppressive therapy
      iv. Chimeric antigen receptor-T-cell therapy or hematopoietic stem cell transplant (within 2 years of transplantation or taking immunosuppressive therapy)
      v. Moderate or severe primary immunodeficiency
      vi. Advanced HIV infection (CD4 cell count <350 cells/$mm^3$)
      vii. Taking high-dose corticosteroids (≥20 mg of prednisone or equivalent per day when administered for at least 2 weeks), B-cell-depleting agents (within the past year), alkylating agents, antimetabolites, transplant-related immunosuppressive drugs, tumor necrosis factor blockers, or other immunosuppressive or immunomodulatory biologic agents

    b. For cohort B, is at risk of acquiring SARS-CoV-2 due to regular unmasked face-to-face interactions in indoor settings (eg, workplace, gym facility, public transportation, etc)

***Note***: Participants who meet criteria 4a and 4b will be enrolled in cohort A.

5. Agrees to defer receipt of any COVID-19 vaccination or booster for a minimum of 28 days after dosing on day 1
6. Provides written documentation of informed consent by signing a current independent ethics committee/institutional review board-approved informed consent form at the time of screening. A legally authorized representative may be used in cases in which inclusion criterion 8 is able to be fulfilled. In the case of adolescents, informed consent/assent must also be obtained as required by local guidelines
7. Has access to a device (eg, mobile phone, tablet, etc) enabled to receive study reminders (eg, SMS text messages)
8. Is able to understand and comply with study requirements/procedures (if applicable, with assistance by a caregiver, surrogate, or legally authorized representative) based on the assessment of the investigator
9. For participants assigned female sex at birth:
    a. Is not of childbearing potential, **OR**
    b. Is of childbearing potential and practicing adequate contraception for at least 28 days before dosing on day 1 through 6 months after any dosing and has a negative pregnancy test result on day 1

***Note***: Pregnant participants will be eligible for enrollment after independent data monitoring committee (IDMC) review of safety lead-in data only upon sponsor communication to sites and only in regions permitted by local health authorities and local ethics committees. If pregnant participants are eligible for enrollment, this criterion is no longer applicable.

*Exclusion Criteria*

Participants are excluded from the study if any of the following criteria apply:

1. For cohort B: prior receipt of a COVID-19 vaccine or booster within 120 days before randomization
2. Prior receipt of convalescent plasma or a monoclonal antibody to SARS-CoV-2 active against currently circulating variants, including in the setting of a clinical trial, within 120 days before randomization
3. Prior known or suspected SARS-CoV-2 infection within 120 days before randomization
4. Exposure to someone with known or suspected SARS-CoV-2 infection in the 5 days before randomization
5. Is acutely ill or has any of the following symptoms suggestive of infection, in the opinion of the investigator:
    - Fever ≥38 °C (≥100.4 °F)
    - Shortness of breath/difficulty breathing
    - Chills (shivering)
    - Cough
    - Fatigue (low energy or tiredness)
    - Muscle or body aches
    - Headache
    - Loss of taste
    - Loss of smell
    - Sore throat
    - Congestion (stuffy or runny nose)
    - Nausea
    - Vomiting
    - Diarrhea
6. Received any investigational product within 30 days or 5 half-lives (whichever is longer) before the day of enrollment
7. Received or plans to receive a non-COVID-19 vaccine within 28 days before or after dosing on day 1 (except for seasonal influenza vaccine, which is not permitted within 14 days before or after dosing on day 1)
8. Known allergy/sensitivity or hypersensitivity to the study drug, including excipients

9. Pregnant, as confirmed with a positive pregnancy test on the day of dosing (day 1), or breastfeeding

   ***Note***: Pregnant/breastfeeding participants will be eligible for enrollment after IDMC review of safety lead-in data only upon sponsor communication to sites and only in regions permitted by local health authorities and local ethics committees. If pregnant participants are eligible for enrollment, this criterion is no longer applicable.
10. Known clinically significant bleeding disorder (eg, factor deficiency, coagulopathy, platelet disorder) or prior history of significant bleeding or bruising following venipuncture. Abnormal coagulation labs or use of anticoagulant medication are not exclusionary in the absence of clinical findings
11. Any serious concomitant systemic disease, condition, or disorder that, in the opinion of the investigator, may lead to hospitalization or death within the study period, confound the results of the study, or confer an additional risk to the participant by their participation in the study
12. Is or has an immediate family member (eg, spouse, sibling, child, guardian/legally authorized representative, parent) who is an investigator or site/sponsor employee directly involved with the study

**PHARMACOKINETICS**

Pemivibart half-life and serum concentration-time profiles were assessed using population pharmacokinetic (popPK) modeling and simulation. Pemivibart serum concentration by timepoint was evaluated for all participants who received any amount of pemivibart during the study and had at least 1 quantifiable serum concentration post dose (PK analysis set). In cohort A, blood PK samples were collected from all participants on days 1 (post dose), 14, and 28 and months 3 (pre and post dose) and 6. In cohort B, blood PK samples were collected from all participants on day 28, and months 3 (pre and post dose) and 6; on days 1 and 10, samples were collected only for a subset of participants. Serum concentrations of pemivibart were assessed using a validated electrochemiluminescence method at PPD® Laboratories (Richmond, VA). An interim pemivibart popPK model was developed using available PK data from a phase 1 study (24 healthy adults; NCT05791318) through month 12 and the CANOPY study (603 adults) from both cohorts through month 6.

**IMMUNOGENICITY**

Immunogenicity was assessed at PPD® Laboratories (Richmond, VA) using a validated electrochemiluminescence method designed to detect antidrug antibodies (ADAs) to pemivibart. In both cohorts, blood ADA samples were collected from all participants on days 1 (pre dose) and 28 and months 3 (pre dose) and 6. ADA was assessed in all participants receiving pemivibart and approximately 10% of participants receiving placebo. If a participant had a negative ADA measurement at baseline and positive ADA post baseline, or a positive ADA measurement at baseline and a positive ADA measurement post baseline with an ADA titer ≥4 times the baseline ADA titer, this is defined as a treatment-emergent (TE) ADA. The minimum required dilution (MRD) of the assay was determined to be 12.5. ADA titers <12.5 (or <MRD) were imputed to 12.5 for the determination of treatment emergent ADA and used to calculate the fold-rise from baseline. Fold-rise from baseline was calculated only for participants with baseline positive ADA. The immunogenicity analysis population included participants who received any amount of study drug and had a valid immunogenicity test result before the initial dose of study drug and at least 1 valid result post dose

**SAMPLE SIZE**

For cohort A, approximately 300 participants were planned to be enrolled. The sample size was originally selected empirically for evaluation of safety and tolerability and to provide meaningful estimates of serum virus neutralizing antibody (sVNA) titers, PK, and immunogenicity in this population and subgroups of interest. For the primary immunobridging analysis of calculated sVNA titers at day 28, a minimum sample size of 200 provides approximately 90% power to test noninferiority with a margin of 80% at 1-sided alpha 0.05 assuming the expected geometric mean ratio is 0.87 with a variability of 40% (coefficient of variation). For cohort B, approximately 450 participants were planned to be randomized in a 2:1 ratio to receive pemivibart (n≈300) or placebo (n≈150). For evaluation of safety, the pooled sample size of cohorts A and B with approximately 600 participants exposed to pemivibart will allow detecting rare events during the study period (ie, detecting an adverse event with a rate ≥0.005 with 95% probability). A placebo control was included in cohort B to allow a robust evaluation of common adverse events.

### SUPPLEMENTARY RESULTS

#### ANAPHYLAXIS EVENTS

Anaphylaxis was reported in 4 participants in cohort A: 2 non-serious events at the initial dose of pemivibart and 2 serious, life-threatening events at redosing at month 3. The 2 cases occurring with the first dose were reported as moderate events of hypersensitivity and infusion-related reactions but were reclassified as anaphylaxis per Sampson's criteria [1] during the regulatory review. One participant experienced flushing, dizziness, tinnitus, and wheezing within 4 minutes of dosing, while the other experienced dyspnea, diaphoresis, face erythema, chest discomfort, and tachycardia immediately after dosing was initiated. Both participants received oral diphenhydramine; the second participant also received inhaled albuterol. The symptoms resolved within 10 minutes and 3 hours of dosing, respectively.

The 2 anaphylaxis cases occurring with the second dose of pemivibart were reported as serious and life threatening. Symptoms included pruritus, urticaria, angioedema, dyspnea, and either erythema or flushing. One of the 2 participants with serious anaphylaxis also experienced headache, dizziness, and chest pain; additionally, pruritus, erythema, and urticaria reoccurred in this participant within 24 hours of the initial onset of anaphylaxis. Both participants were treated with diphenhydramine and epinephrine, and 1 participant also received oral prednisone and metoprolol for an associated flare of an underlying condition. The symptoms resolved within the same day of dosing with no sequelae (~5 hours and 20 minutes later) and next day of dosing with sequelae (flare of underlying condition), respectively.

#### PopPK MODEL COVARIATE ASSESSMENTS

The popPK model covariate assessment showed no systematic difference in PK between participants with or without immune compromise [2]. The PK of pemivibart was not affected by age, race, or obesity. Body weight is related to the variability in PK such that heavier adults are predicted to have lower exposure, but it is not expected to have a clinically relevant effect on the PK of pemivibart through month 3 post dose in individuals with body weights ranging from 43 to 190 kg. The relationship identified between sex and clearance or volume of the central compartment is also not expected to be clinically relevant.

**IMMUNOGENICITY**

In cohort A, 7 (2.3%) participants were confirmed positive for ADAs at baseline (day 1 pre dose); of these, 5 had a predose titer at the minimum required dilution or less (≤12.5). Treatment-emergent ADAs were observed in 6 (2.0%) participants, including 2 (0.7%) at day 28, 2 (0.7%) at day 90, and 4 (1.3%) at day 180, all with titers ≤12.5.

In cohort B, 1 (0.3%) participant was confirmed positive for ADAs at baseline. TE ADAs were observed in 3 (1%) participants, and only at day 180 with titers ≤12.5.

In both cohorts, TE ADAs were not observed in participants confirmed positive for ADAs at baseline.

### SUPPLEMENTARY TABLE

#### Supplementary Table 1. TEAEs Leading to Study Treatment Discontinuation (Cohorts A and B, Safety Analysis Set)

| Parameter, n (%) | Cohort A | |
|---|---|---|
| | Pemivibart n=306 | |
| **Any study treatment discontinuation[a]** | 12 (3.9) | |
| **TEAEs leading to study treatment discontinuation** | 7 (2.3) | |
| **Study-drug related** | 7 (2.3) | |
| Anaphylactic reaction | 2 (0.7) | |
| Hypersensitivity | 1 (0.3) | |
| Infusion related hypersensitivity reaction | 1 (0.3) | |
| Infusion related reaction | 2 (0.7) | |
| Tachycardia | 1 (0.3) | |
| Tremor | 1 (0.3) | |
| **Parameter, n (%)** | **Cohort B** | |
| | **Pemivibart n=317** | **Placebo n=162** |
| **Any study treatment discontinuation[a]** | 23 (7.3) | 8 (4.9) |
| **TEAEs leading to study treatment discontinuation** | 5 (1.6) | 0 |
| **Study-drug related** | 3 (0.9) | 0 |
| Hypersensitivity | 2 (0.6) | 0 |
| Eye pain | 1 (0.3) | 0 |
| **Study-drug unrelated** | 2 (0.6) | 0 |
| Bradycardia | 1 (0.3) | 0 |
| Ventricular fibrillation | 1 (0.3) | 0 |

[a]A participant was considered to have discontinued treatment if they did not receive the second dose or only received a partial amount of the second dose.

### SUPPLEMENTARY REFERENCES


1. Sampson HA, Muñoz-Furlong A, Campbell RL, et al. Second symposium on the definition and management of anaphylaxis: summary report--Second National Institute of Allergy and Infectious Disease/Food Allergy and Anaphylaxis Network symposium. J Allergy Clin Immunol **2006**; 117: 391-7.
2. Rubino C, Cammarata A, DeRyke A, et al. Population pharmacokinetics of pemivibart (VYD222), an extended–half-life monoclonal antibody in development for the pre-exposure prophylaxis of COVID-19. Presented at: MAD-ID 2024; 8-11 May 2024; Orlando, Florida.
